# Trends of attrition from HIV care and its predictors among Adolescent Girls and Young Women with inconsistent viral load suppression results in Mainland Tanzania, 2016–2024

**DOI:** 10.1101/2025.11.04.25339531

**Authors:** Anthony Charles Kavindi, Jegede Feyisayo Ebeneezer, Asteria Karungi Nyongoli, Nagalal William, Paschal Yuda, Deogratius W. Kinoko, Laura J. Shirima, Festo Charles, Marion Sumari-de Boer

**Affiliations:** School of Public Health, KCMC University, P.O. Box 2240, Kilimanjaro Tanzania; Kilimanjaro Clinical Research Institute; Management and Development for Health (MDH); Bayero University Kano, Department of Life Science, Nigeria

## Abstract

**Background:** Adolescent Girls and Young Women (AGYW) in Tanzania Mainland are disproportionately affected by HIV and are at high risk of attrition from HIV care, undermining efforts to achieve the UNAIDS 95-95-95 targets by 2030. AGYW with inconsistent viral load suppression are particularly vulnerable, leading to suboptimal treatment outcomes and continued transmission risks.

**Methods:** A retrospective cohort study was conducted using data from the national CTC2 database. Survival analysis techniques were employed to estimate attrition incidence rates and identify predictors among AGYW with inconsistent viral load suppression results over a five-year follow-up period.

**Results:** The overall incidence rate of attrition was 11.8 per 1,000 person-years. Higher attrition was observed among AGYW aged 20–24 years (AHR: 1.58) compared to those aged 15-19 years, those residing in rural areas (AHR: 1.15) compared to those residing on urban area, with initial viral load ≥1,000 cp/mL (AHR: 1.28) compared to those with viral load < 1000 cp/mL, and those attending public health facilities (AHR: 1.79) compared to private or FBO facilities. Protective factors included being on second-line ART (AHR: 0.63), longer ART duration ≥4 years (AHR: 0.43), and residence in the Lake Zone (AHR: 0.64).

**Conclusion:** Early attrition from HIV care is common among AGYW with inconsistent viral load suppression, particularly in the first year. Tailored interventions targeting at-risk groups—based on age, residence, ART regimen, and facility type are urgently needed to improve retention and treatment outcomes in this population.

## Introduction

Human Immunodeficiency Virus (HIV) is a virus that weakens the immune system by targeting and destroying CD4 T cells, which play a crucial role in defending the body against infections. Without treatment, HIV progresses through four stages: acute infection, clinical latency, symptomatic HIV infection, and finally, acquired immunodeficiency syndrome (AIDS) (1).

Globally, HIV remains a major public health concern. As of 2024, approximately 39.9 million people were living with HIV (2). Adolescent Girls and Young Women (AGYW) account for 1.9 million of these cases, and about 4,000 AGYW are newly infected each week. Alarmingly, 3,100 of these new infections occur in Sub-Saharan Africa (SSA), accounting for 77% of all new AGYW infections globally in 2023, with Eastern and Southern Africa alone contributing around 60% (2).

In response, UNAIDS launched the 95-95-95 targets aimed at ending the AIDS epidemic by 2030: 95% of people living with HIV should know their status, 95% of those diagnosed should be on antiretroviral therapy (ART), and 95% of those on ART should achieve viral suppression (UNAIDS, 2023). However, by 2023, global achievements still lagged: 86% of people living with HIV knew their status, 77% were on ART, and 72% were virally suppressed (3).

Effective HIV prevention strategies, including condom use, pre-exposure prophylaxis (PrEP), and prevention of mother-to-child transmission (PMTCT), are essential to reducing new infections (4). Moreover, ensuring widespread ART coverage leads to viral suppression, reducing the risk of HIV transmission (5)Engagement of communities and stigma reduction efforts are also crucial to expanding access to ART for key populations, such as AGYW, sex workers, and men who have sex with men (1).

Despite progress, retention in HIV care remains a major challenge in SSA. A study by Haas et al. (2018) found that five years after ART initiation, only 52.1% of patients were retained in care, with 41.8% lost to follow-up and 6.0% deceased (6). When accounting for undocumented deaths and self-transfers, the retention estimate increased to 66.6%. Similarly, a meta-analysis of 32 ART programs in SSA reported decreasing retention rates over time: 80% after one year, 77% after two years, and 72% after three years, with loss to follow-up and death being the main contributors to attrition (7).

In Tanzania, the 2023 Tanzania HIV Impact Survey (THIS) reported an HIV prevalence of 4.5% among adults, with higher rates in women (5.6%) than in men (3.0%) (8).Among AGYW aged 20–24 years, the prevalence was 1.8%, double that of males in the same age group (0.9%). The HIV incidence among adults was 0.18%, while AGYW had a higher incidence of 0.33%, contributing 80% of new infections (8).

While adult PLHIV on ART showed a viral suppression rate of 94.3% (94.9% among women and 92.9% among men), the rate was notably lower among AGYW at 86.6%. Even more concerning, among all AGYW living with HIV (regardless of ART status), only 51.3% were virally suppressed, compared to 71.5% among young men (8). These figures suggest that AGYW face heightened risks of disease progression and ongoing transmission due to inconsistent viral suppression and retention in care.

Understanding the patterns and predictors of attrition from HIV care among AGYW with unsuppressed or inconsistent viral load is therefore critical to achieving HIV epidemic control. This study seeks to address this gap by exploring trends in ART care attrition, survival time in care, and associated factors among AGYW in Tanzania Mainland.

## Methods

### Data source and study design

This study employed a retrospective cohort design utilizing routinely collected, de-identified data of people living with HIV (PLHIV) attending Care and Treatment Clinics (CTCs) across Mainland Tanzania from 2016 to 2024. The data, collected by healthcare providers at over 1,100 health facilities, include socio-demographic, clinical, laboratory, and pharmacological information documented during patient visits.

The study population comprised adolescent girls and young women (AGYW) aged 15–24 years with documented inconsistent HIV viral load suppression, defined as having at least two viral load results above 1000 copies/mL after six months on ART. Participants enrolled between 2016 and 2019 were followed for up to five years to assess attrition from HIV care and associated factors.

Data were extracted from the CTC2 database, which systematically collects patient data through linked patient-held CTC1 cards and facility-based CTC2 cards, supported by pre-ART and ART registers and cohort analysis reports. The unique patient identification numbers ensured consistent longitudinal tracking.

Eligibility criteria included AGYW aged 15–24 years at enrollment with at least two documented high viral load test results. Exclusions were patients transferred without follow-up data or lacking viral load tests during the study period. From an initial pool of over 2.6 million PLHIV, 7,910 eligible AGYW with inconsistent viral load suppression were identified and included in the final analysis.

### Study Variables

#### Dependent Variable

The dependent variable in this study was attrition from HIV care. It was defined as a binary outcome; Yes (1) for individuals who had ever experienced attrition (i.e., were not in care at any point during the follow-up period) and No (0) for those who remained continuously engaged in HIV care throughout the follow-up period.

#### Independent Variables

Independent (predictor) variables were categorized into socio-demographic, clinical, and health system factors (Table 1)

**Table 1.** Categorization and coding of independent variables.

| **Variable** | **Definition** | **Coding** |
| --- | --- | --- |
| Age at ART initiation | Age calculated from date of birth to date of ART start | 15–19 years, 20–24 years |
| Age at study enrollment | Age of the client at the time of study enrollment | 0–14 years, 15–19 years, 20–24 years |
| Residence | Geographical location of the client | Urban, Rural |
| Marital status | Self-reported marital status from CTC2 database | Single, Married, Divorced/Separated |
| Type/Level of health facility | Healthcare facility hierarchy based on service complexity | Dispensary, Health Center, Hospital |
| Health facility ownership | Ownership and management type of healthcare facility | Government, Private, Faith-Based Organization |
| Zone | Zone of Mainland Tanzania based on geographical regions | Coastal, Northern Highland, Lake, Central, Southern Highland, Western |
| Entry point at HIV diagnosis | Location/service point where HIV diagnosis was made | OPD/IPD, RCH/PMTCT, VCT, Community, Others |
| Initial viral load | First viral load after 6 months of ART, in copies/mL | <50 (Undetectable), 50–999 (LLV), ≥1000 (Unsuppressed) |
| Baseline CD4+ cell count | CD4 count measured at ART initiation using flow cytometry | <200, 200–500, >500 cells/mL |
| Baseline HIV disease stage | WHO clinical staging (I–IV) at ART enrollment | Stage I, II, III, IV |
| Duration on ART | Time on ART from initiation to study endpoint | <4 years, ≥4 years |

#### Data Analysis Plan

Analyses were performed using Stata version 18.0 (StataCorp LLC, College Station, TX, USA). Descriptive statistics summarized participants’ sociodemographic and clinical characteristics. Continuous variables (e.g., age at enrollment, age at ART initiation, and duration on ART) were summarized using medians and interquartile ranges (IQRs), while categorical variables were summarized using frequencies and proportions.

For Objective 1, which examined five-year trends in attrition from HIV care among adolescent girls and young women (AGYW) with inconsistent viral load suppression, time-to-event data were prepared using the *stset* command. Attrition rates per 1,000 person-years were estimated for each follow-up year using *strate*, and temporal trends were visualized with line graphs.

For Objective 2, Kaplan–Meier survival analysis estimated median survival time and cumulative retention probabilities. Time-to-event was defined from ART initiation to attrition (loss to follow-up, treatment cessation, or death). Survival curves were stratified by marital status, duration on ART, ART regimen, and initial viral load, with differences assessed using the log-rank test.

For Objective 3, predictors of attrition were assessed using a Weibull regression model with gamma-distributed shared frailty at the individual (CTC ID) level. Univariable models were fitted first, and variables with likelihood ratio test p-values <0.2 were included in the multivariable model. Multicollinearity was checked using the Variance Inflation Factor (VIF), and model fit was assessed via likelihood ratio tests. Confounding was evaluated by comparing crude and adjusted hazard ratios, with a ≥15% change indicating potential confounding.

#### Ethical Considerations

Ethical approval was obtained from the KCMC University Research Ethics Review Committee (CRERC) (Approval ID: PG195/2024, dated 7 March 2025). Data access permission was granted by the National AIDS and STI Control Programme (NASCOP), Ministry of Health, Tanzania (Reference No. PA.104/262/01/51, dated 26 May 2025). The study used anonymized secondary data; thus, informed consent was not required. Confidentiality was maintained by excluding all personal identifiers from the analysis and reporting.

## Results

### Socio-demographic and clinical characteristics

A total of 7,910 AGYW were analyzed. The median age at ART initiation was 17 years (IQR: 9– 18), with 65.0% starting before age 15. At enrollment, the median age was 17 years (IQR: 15–21), and 66.7% were aged 15–19 years. Among those with known marital status (n=6,256), 76.4% were single and 22.2% married. Most received care at public facilities (71.6%) and hospitals (53.4%), while 25.5% attended faith-based facilities. HIV was mainly diagnosed at VCT (51.4%), followed by OPD/IPD (25.0%) and PMTCT/RCH (13.8%) (n=7,141). Most participants were from the Southern Highlands (30.6%) and Coastal Zones (26.9%), with 51.7% in urban areas. First-line ART regimens were used by 90.4%, with a median duration of 105 months (IQR: 83–141). At ART initiation, 30.0% had CD4 <200 cells/mm³ and 32.9% ≥500. Over half (52.3%) were WHO Stage 3, and initial viral load showed 56.4% unsuppressed (≥1000 copies/mL) and 23.9% undetectable (<50 copies/mL) (See Table 1).

**Table 1:** Social Demographic and clinical characteristics of the study AGYW (n=7,910).

| Variable | Frequency (n=7,910) | Percentage (%) |
| --- | --- | --- |
| Age at start ART |  |  |
| 0-14 | 5,142 | 65.0 |
| 15-19 | 1,460 | 18.5 |
| 20-24 | 1,308 | 16.5 |
| *Median (IQR)* | 17(9,18) |  |
| **Age at study enrollment** |  |  |
| 15-19 | 5,274 | 66.7 |
| 20-24 | 2,636 | 33.3 |
| *Median (IQR)* | 17(15,21) |  |
| **Marital Status (n=6,256) \*** |  |  |
| Single | 4,780 | 76.4 |
| Married | 1,386 | 22.2 |
| Divorced | 90 | 1.4 |
| **Facility ownership** |  |  |
| Private | 234 | 3.0 |
| Public | 5,662 | 71.6 |
| FBO | 2,014 | 25.5 |
| **Facility type** |  |  |
| Dispensary | 1,297 | 16.4 |
| Health Centre | 2,285 | 28.9 |
| Hospital | 4,225 | 53.4 |
| Other | 103 | 1.3 |
| **Entry point at HIV diagnosis (n=7,141) \*** |  |  |
| OPD/IPD | 1,786 | 25.0 |
| RCH/PMTCT | 988 | 13.8 |
| VCT | 3,673 | 51.4 |
| COMMUNITY | 124 | 1.7 |
| OTHERS | 570 | 8.0 |
| **Zone** |  |  |
| Central Zone | 550 | 7.0 |
| Coastal Zone | 2,129 | 26.9 |
| Lake Zone | 1,371 | 17.3 |
| Northern Zone | 1,247 | 15.8 |
| Southern Highland Zone | 2,420 | 30.6 |
| Western Zone | 193 | 2.4 |
| **Residence** |  |  |
| Urban | 4,091 | 51.7 |
| Rural | 3,819 | 48.3 |
| **ART regimen** |  |  |
| First Line | 7,148 | 90.4 |
| Second line | 762 | 9.6 |
| **CD4 4-at baseline (n=6,868) \*** |  |  |
| <200 | 2,063 | 30.0 |
| 200 - 349 | 1,426 | 20.8 |
| 350 - 499 | 1,117 | 16.3 |
| 500+ | 2,262 | 32.9 |
| **WHO stage (n=7,578) \*** |  |  |
| Stage 1 | 1,222 | 16.1 |
| Stage 2 | 1,277 | 16.9 |
| Stage 3 | 3,966 | 52.3 |
| Stage 4 | 1,113 | 14.7 |
| **Duration on ART** |  |  |
| <3 | 89 | 1.1 |
| 4+ | 7,821 | 98.9 |
| *Median (IQR)* | 105(83,141) |  |
| **Initial VL result** |  |  |
| <50 undetectable | 1,889 | 23.9 |
| 50-999 LLV | 1,556 | 19.7 |
| >=1000 unsuppressed | 4,465 | 56.4 |

Key: \* Frequencies (n) do not tally to the total due to the missing value; OPD: Outpatient Department VCT: voluntary Counseling and Testing, IPD:
Inpatient Department, RCH: Reproductive and Child Health, LLV: Low-Level Viremia.

### Overall attrition rate from HIV care

A total of 5,100 attrition events occurred over 432,573.7 person-years (PY) of follow-up, yielding an overall attrition rate of 11.8 per 1,000 PY (95% CI: 11.15–11.21). This implies that for every 1,000 AGYW followed for one year, approximately 11.8 were not retained in care.

### Five-year trends in attrition rates

Attrition was highest in the first year at 15.8 per 1,000 PY (95% CI: 15.1–16.7) and declined over time: 13.1 (year 2), 12.1 (year 3), 8.9 (year 4), and 7.4 per 1,000 PY (95% CI: 6.8–8.1) in year 5 (Figure 1).

**Fig 1.**
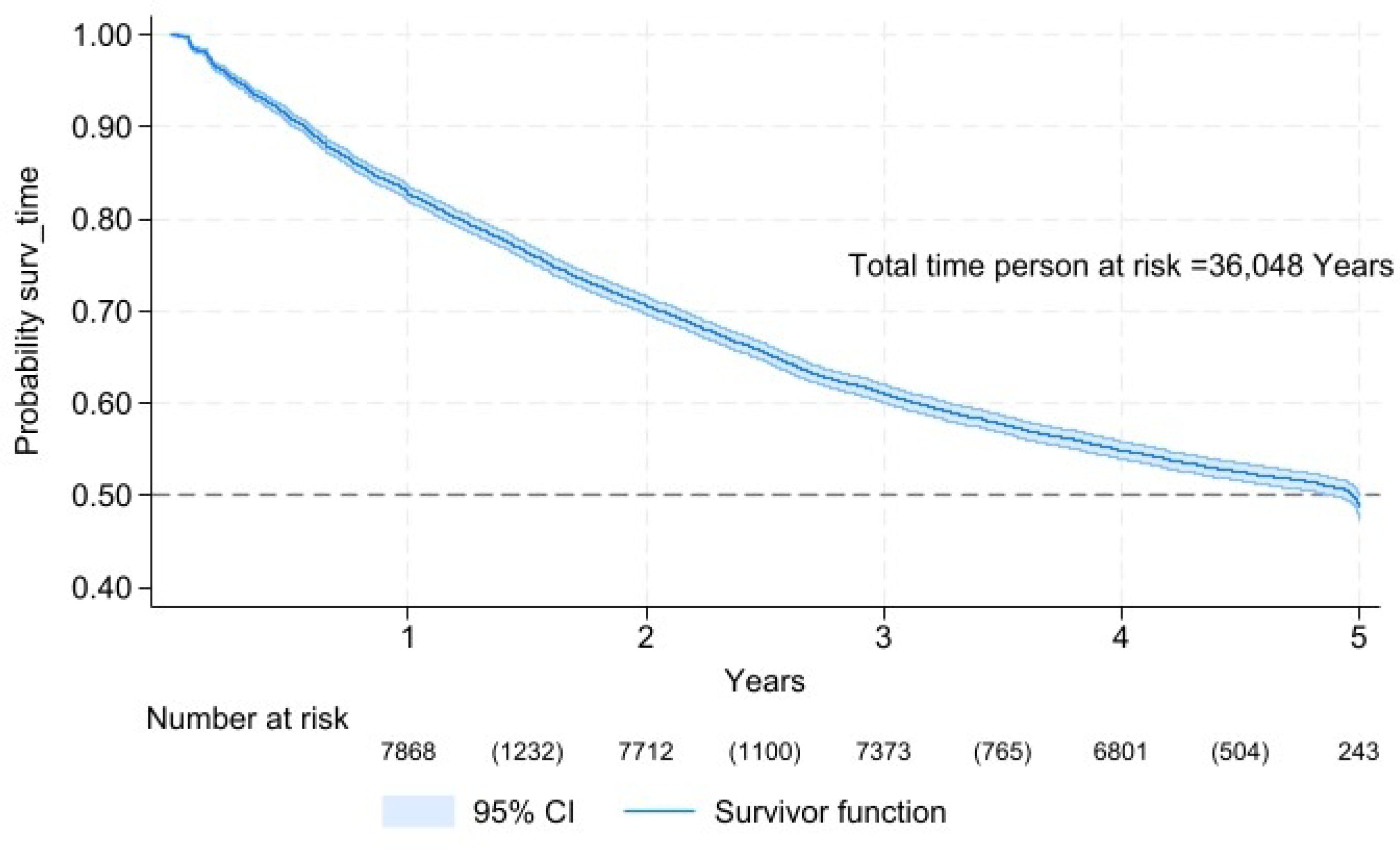
Flow chart showing the recruitment of the AGYW with inconsistent viral load suppression.

**Fig 2.**
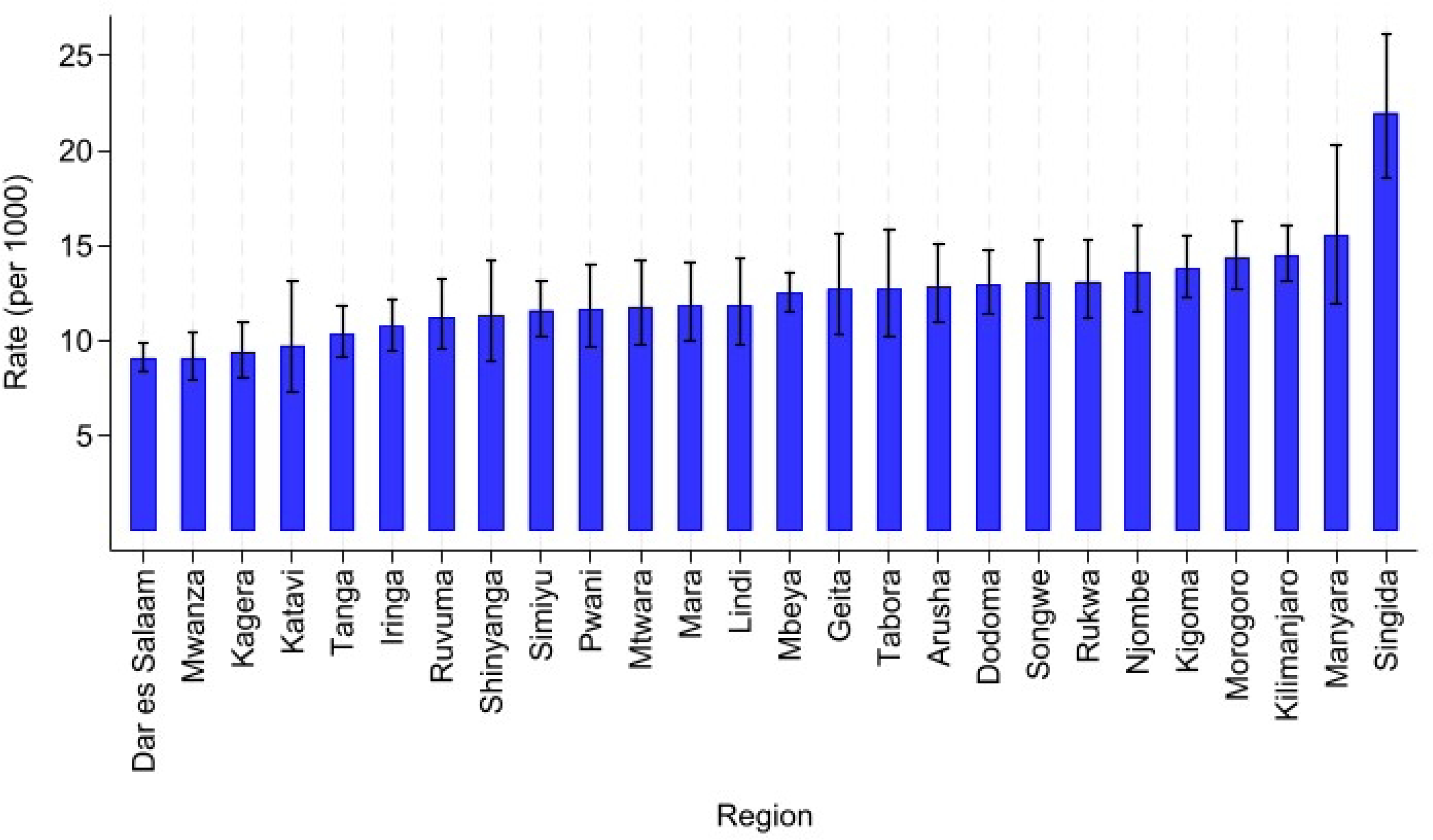
Five-year follow up trend of attrition rates from HIV care over time per 1000 years.

### Attrition rate from HIV care among AGYW by geographical zones

The analysis revealed notable variations in attrition rates from HIV care among adolescent girls and young women (AGYW) with inconsistent viral load suppression across geographical zones in Mainland Tanzania. As illustrated in Figure 4.3, the Central Zone exhibited the highest attrition rate, with 14.78 per 1,000 person-years (PY) (95% CI: 13.46–16.22), indicating a comparatively elevated risk of disengagement from care. This was followed by the Northern Zone (12.91 per 1,000 PY), Southern Highland Zone (12.42 per 1,000 PY), and Western Zone (11.48 per 1,000 PY). Conversely, the Coastal Zone and Lake Zone demonstrated the lowest attrition rates, at 10.57 per 1,000 PY (95% CI: 10.00–11.19) and 10.41 per 1,000 PY (95% CI: 9.71–11.15), respectively, suggesting relatively stronger retention in care within these zones.

Figure 3 presents the attrition rates from HIV care among adolescent girls and young women (AGYW) with inconsistent viral load suppression across regions in Mainland Tanzania, expressed per 1,000 person-years (PY) with corresponding 95% confidence intervals. Singida Region recorded the highest attrition rate at 22.01 per 1,000 PY (95% CI: 18.54–26.12), reflecting a significantly elevated risk of disengagement from care. Other regions with relatively high attrition rates included Manyara, Kilimanjaro, and Morogoro, each exceeding 14 per 1,000 PY. In contrast, Dar es Salaam, Mwanza, Kagera, and Katavi reported the lowest attrition rates, all below 10 per 1,000 PY, with Dar es Salaam demonstrating the most precise estimate (95% CI: 8.41–9.84), indicating better retention in HIV care within the region.

**Fig 3.**
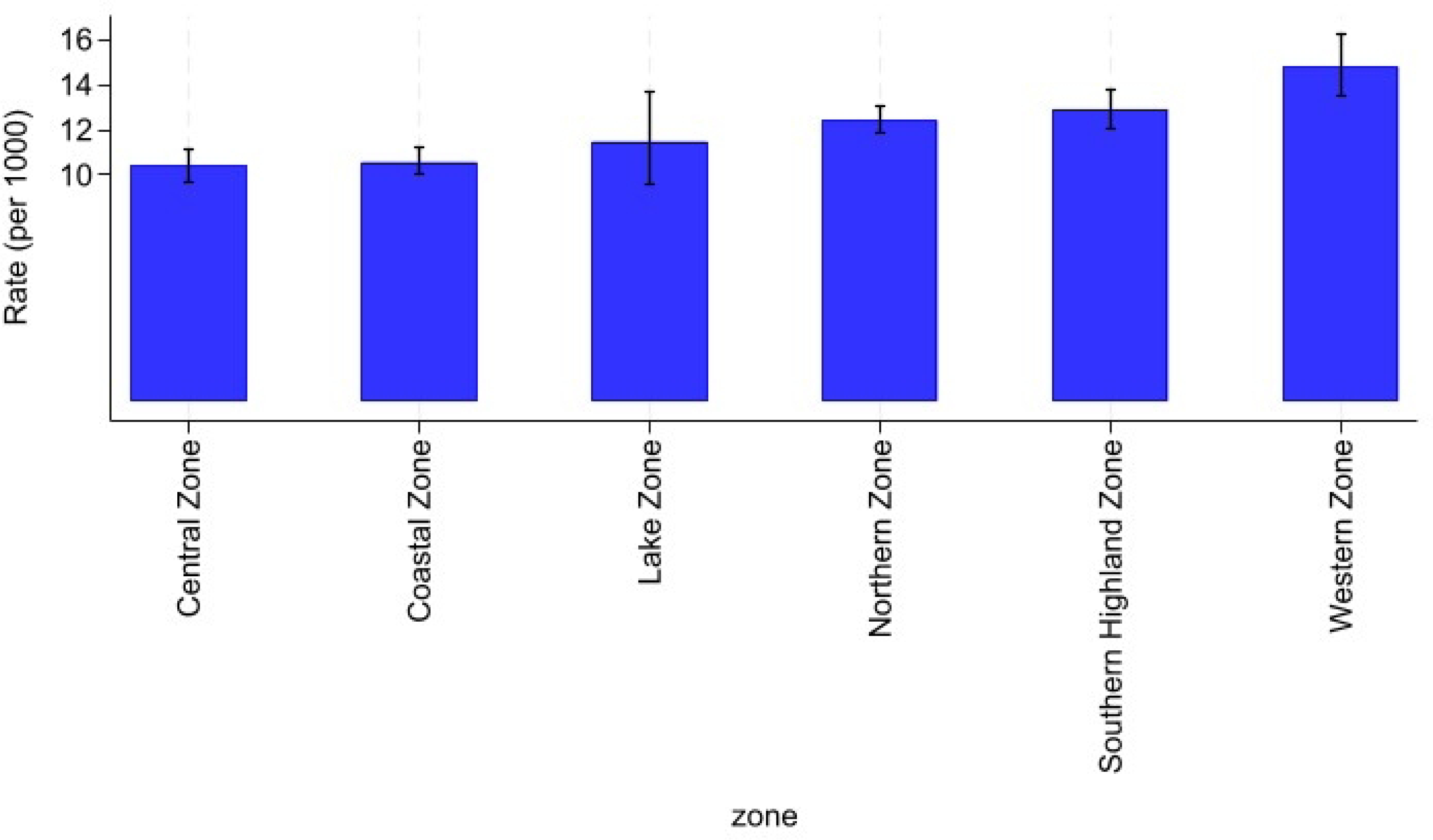
Attrition rate from HIV care among AGYW by geographical zones.

### Survival time in HIV care

To estimate attrition from HIV care among adolescent girls and young women (AGYW) over a five-year follow-up period, survival analysis revealed a progressive decline in retention over time. At the end of the first year, approximately 82.7% of AGYW remained in care, which decreased to 70.6% by the second year, 61.1% by the third year, 54.9% by the fourth year, and 48.7% by the fifth year. These findings indicate that nearly half of the cohort experienced attrition within five years of follow-up. Overall, there was a consistent decline in the probability of remaining in care over time. The median survival time defined as the time by which 50% of the participants were still engaged in care was reached between the fourth and fifth year, suggesting that half of the AGYW cohort remained in HIV care for approximately 4 to 5 years following ART initiation (Figure 4).

**Fig 4.**
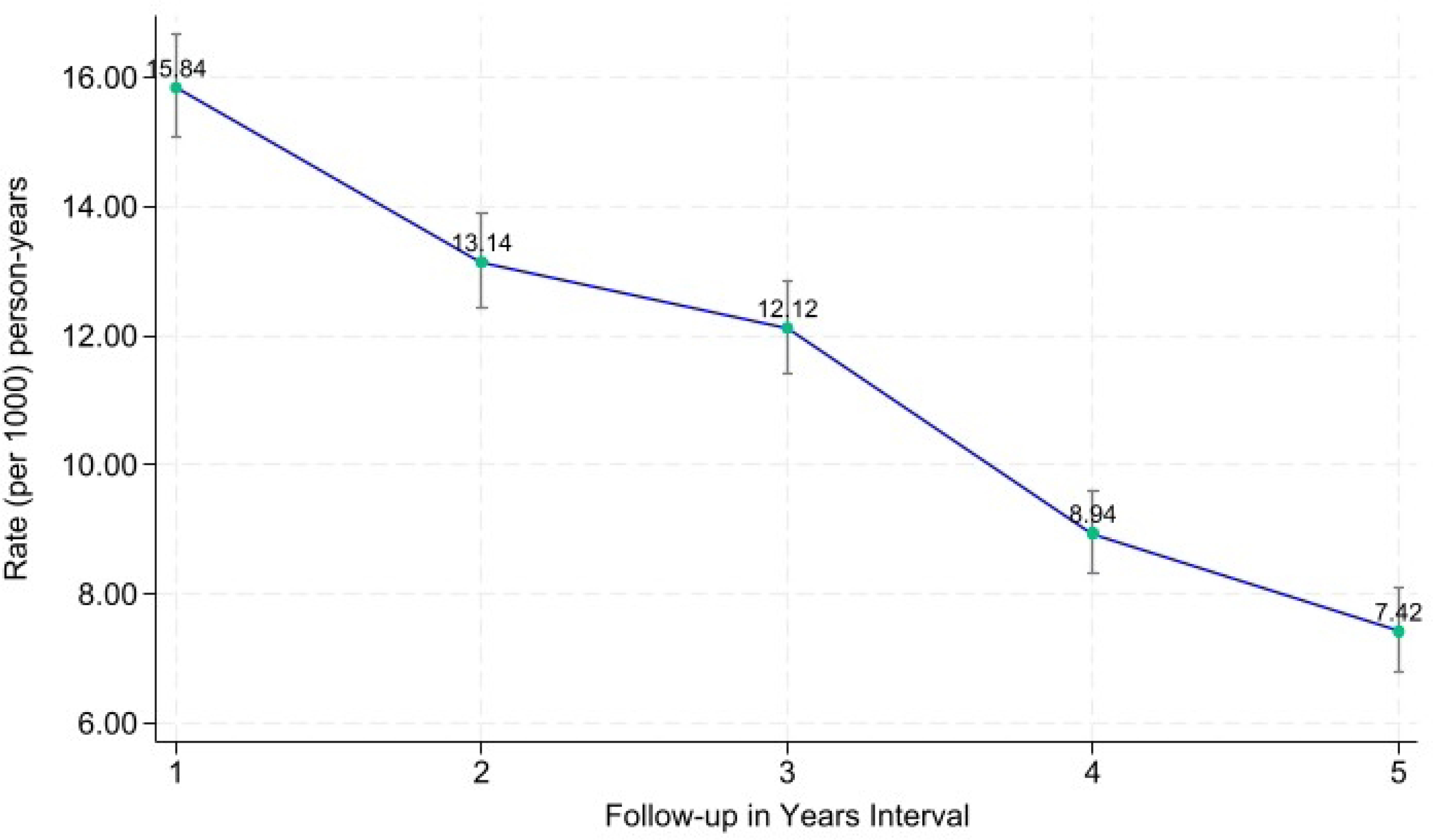
Attrition rate from HIV care among AGYW by Region.

**Fig 5.**
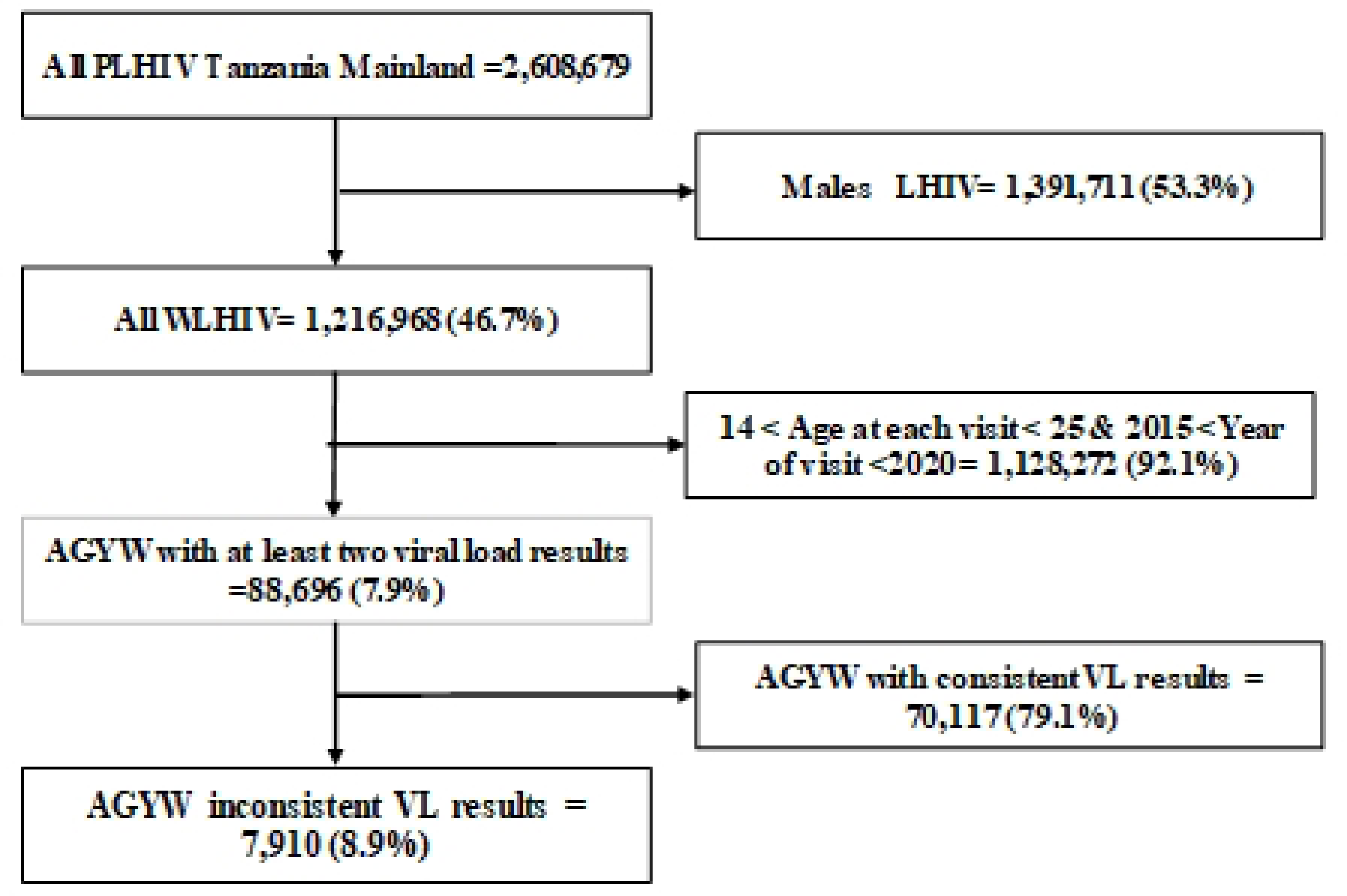
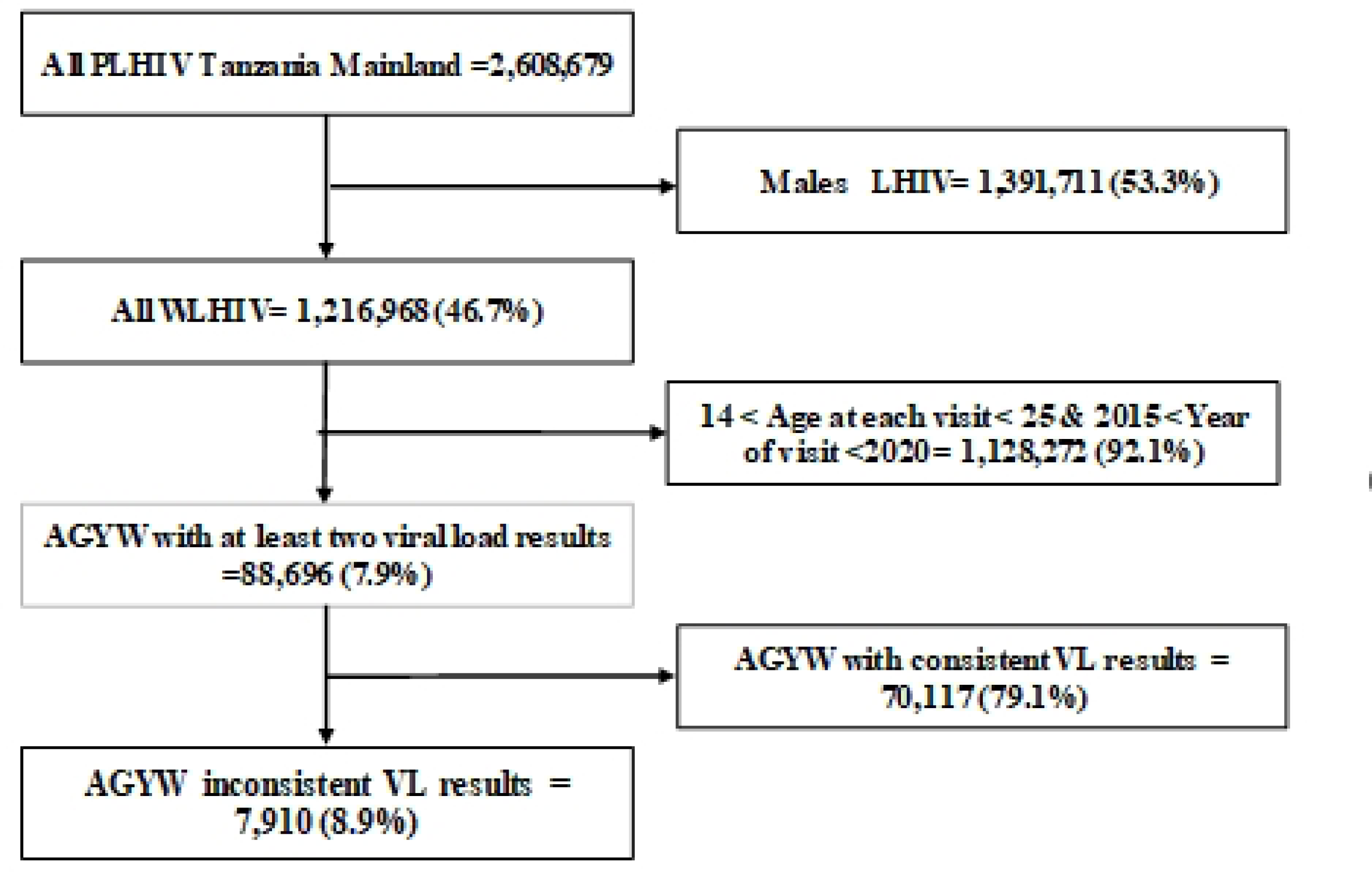
Probability of attrition from HIV care among the AGWY with inconsistent viral load suppression over time.

### Sociodemographic and Clinical Predictors of Attrition from HIV Care

A multivariable Weibull proportional hazards regression model with gamma shared frailty was used to identify predictors of attrition from HIV care among AGYW with inconsistent viral load suppression in mainland Tanzania (2016–2024). Individual-level frailty accounted for unobserved heterogeneity (θ = 0.50; 95% CI: 0.41–0.62; p < 0.001). The Weibull shape parameter was <1 (p = 0.87; 95% CI: 0.84–0.90), indicating a declining hazard over time and higher attrition early in care.

After adjustment, adolescents aged 15–19 and young women aged 20–24 at ART initiation had higher attrition risk than those aged 0–14 (AHR = 1.50 and 1.58; p < 0.001). Conversely, those aged 20–24 at enrollment were less likely to disengage than 15–19-year-olds (AHR = 0.85; p = 0.028). Receiving care from public or faith-based facilities increased attrition risk compared to private facilities (AHR = 1.79 and 1.73), and clients in “Other” facility types had nearly threefold higher risk (AHR = 2.68; p < 0.001).

Regional differences were evident: clients from Coastal, Lake, and Southern Highland Zones had lower attrition than those in the Central Zone. Rural residence increased attrition risk (AHR = 1.15; p = 0.002). Participants with initial viral load ≥1000 copies/mL had 28% higher attrition (AHR = 1.28; p < 0.001). Those on second-line ART (AHR = 0.63; p < 0.001) and those on ART for ≥4 years (AHR = 0.43; p < 0.001) were less likely to disengage. (See Table 2)

**Table 2.**
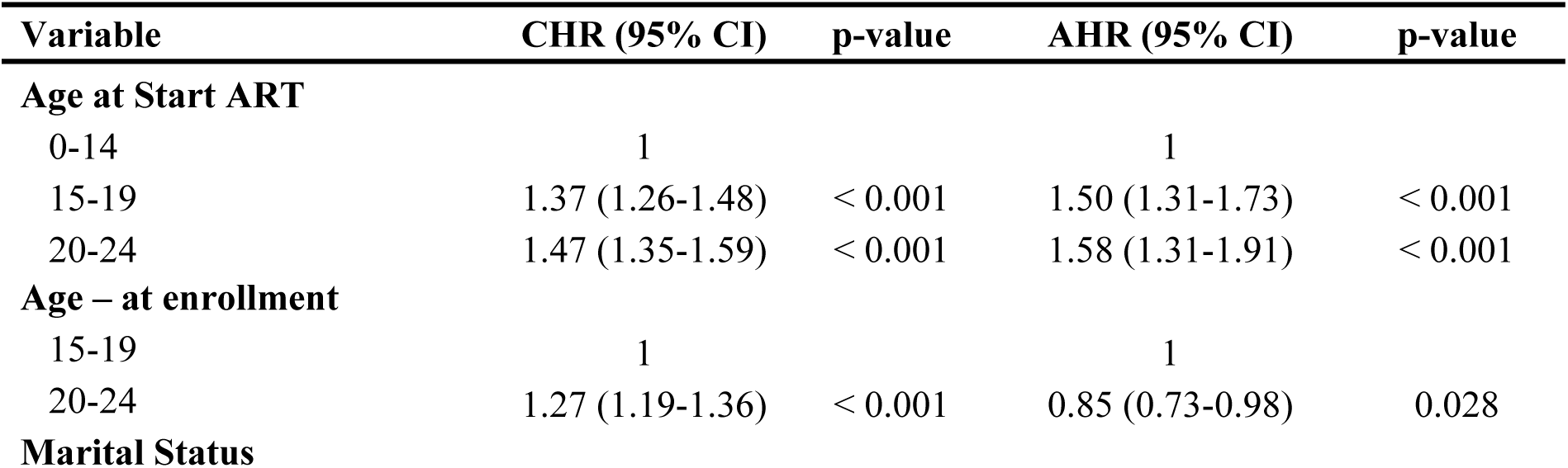
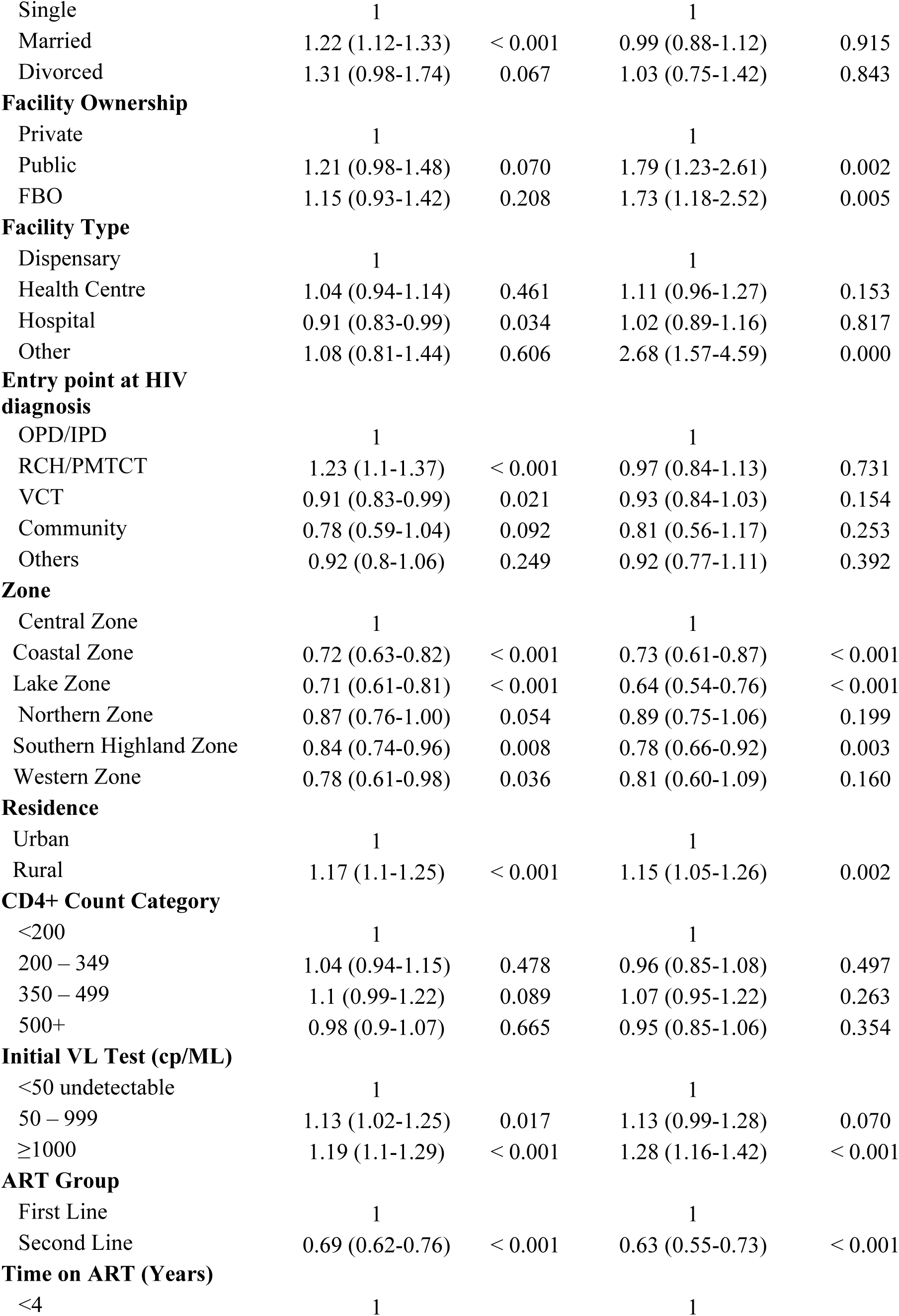
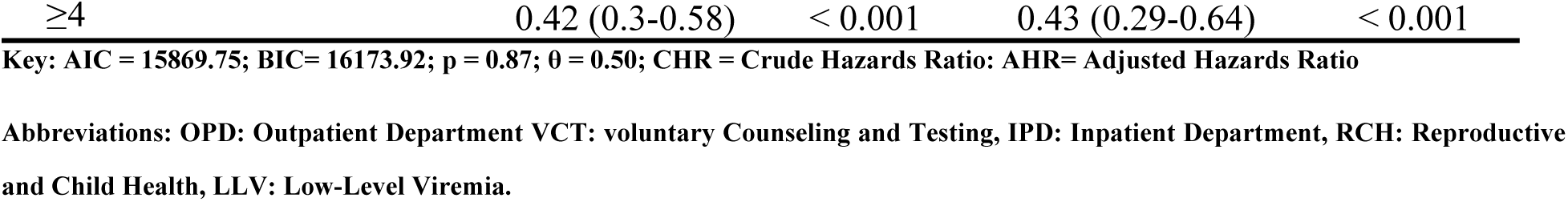
Multivariable analysis for social demographic and clinical predictors of attrition from HIV care among AGYW with inconsistent viral load suppression in Mainland Tanzania from 2016 to 2024 (N= 5,093).

## Discussion

This retrospective cohort study assessed five-year trends and predictors of attrition from HIV care among AGYW with inconsistent viral load suppression in mainland Tanzania (2016–2024), using routine CTC data. The overall attrition rate was 11.8 per 1,000 person-years (PY), aligning with findings from Namibia (9) and sub-Saharan Africa (10), but lower than rates reported in Kenya, Cameroon (11,12), and Mozambique (13). Early attrition was highest during the first year of follow-up, consistent with studies from South Africa, highlighting initial treatment challenges and psychosocial stressors.

Attrition was notably higher among AGYW aged 20–24 compared to younger age groups, echoing regional evidence that transitions to adult responsibilities contribute to disengagement. Those with initial unsuppressed viral loads had a higher risk of attrition, supporting WHO findings that poor viral suppression predicts future disengagement (14).

Attrition declined steadily over the five years, but remained substantial, underlining the need for adolescent-specific interventions like peer support, mobile health (mHealth) strategies, and community outreach. Marital status influenced retention, with married and divorced AGYW experiencing higher attrition than single peers though findings on this vary across settings (15,16)

Clients on first-line ART had higher attrition than those on second-line regimens, likely due to the enhanced monitoring and support often provided to second-line clients (17). Interestingly, attrition decreased with advanced WHO clinical stages possibly due to more intensive care for those with severe disease though this trend contradicts findings in other studies (13).

Entry point into care also impacted outcomes; AGYW diagnosed through PMTCT/RCH and OPD services had higher attrition compared to those entering via VCT or community channels. Younger age at ART initiation (<15 years) was protective, while older initiation ages were associated with higher hazard of attrition, consistent with prior studies in Uganda (15).

Higher attrition was also observed among AGYW with initial viral loads ≥1,000 copies/mL and those residing in rural areas, where geographic and structural barriers may limit consistent access to care. These findings reinforce the importance of tailoring HIV services to adolescent needs, especially for rural and high-risk populations (15,18).

### Strength and limitation of the study

This study has several limitations. Classifying individuals lost to follow-up as attrition may have led to an overestimation of attrition rates, as some clients might have continued care at other facilities under different CTC identification numbers. Additionally, the dataset lacked key behavioral, social, and structural variables such as stigma, mental health status, and household support, which are important predictors of attrition. The use of routine programmatic data collected for service delivery rather than research may also have introduced issues such as incomplete records, misclassification of outcomes, and data inconsistencies due to high workloads and limited data validation at facility level. Despite these limitations, the study has notable strengths. It used a large, nationally representative dataset from the CTC2 database, covering all regions and facilities offering HIV services in Tanzania. The long five-year follow-up period and large sample size increased statistical power, while the use of real-world data enhances the relevance of findings for policy and program decision-making. The focus on adolescent girls and young women—a population disproportionately affected by HIV further adds value to the study by identifying critical gaps in care for this vulnerable group.

## Conclusion

This study found that the overall attrition rate from HIV care among adolescent girls and young women (AGYW) with inconsistent viral load suppression was highest during the first year of follow-up. Notably, individuals with an initial unsuppressed viral load were more likely to disengage from care. Furthermore, the median survival time in HIV care progressively declined over the study period, with the first-year post-enrollment emerging as the most critical period for retention. These findings highlight the urgent need to strengthen early antiretroviral therapy (ART) support interventions during this vulnerable phase. In addition, the study identified several predictors of attrition from HIV care, including older youth (20–24 years), first-line ART regimen, rural residence, high initial viral load (≥1,000 copies/mL), and diagnosis through RCH/PMTCT or OPD services. These results underscore the importance of implementing differentiated service delivery models that promote early viral suppression, expand decentralized care, and provide targeted support for AGYW at elevated risk of disengagement from HIV services.

## Recommendation

To address the high attrition from HIV care among AGYW with inconsistent viral load suppression, an integrated approach combining facility and community-level interventions is essential.

Key strategies include enhancing support in the first year of ART through structured peer mentorship, improved adherence counseling, and digital tools such as mPasha, TIBU, and C3 Tracker for reminders and follow-up. Youth-friendly service models should be reinforced with adolescent-focused clinic days, extended service hours, and mHealth platforms like SMS reminders, telehealth, and mobile apps (e.g., Jichunge and T-HIT), although national coverage remains limited.

In rural areas where attrition is more pronounced efforts should emphasize community ART groups, outreach services, and multi-month dispensing. For clients with initially high viral loads, timely monitoring, intensified adherence counseling, and prompt regimen changes are crucial. Strengthened follow-up systems, particularly for AGYW enrolled through PMTCT and OPD, and involvement of community health workers for home visits and tracing, are also recommended. Lastly, addressing social and marital barriers through disclosure-supportive counseling and partner engagement can improve long-term retention.

## Data Availability

The data used in this study are owned by the National HIV AIDS and STI Control Programme (NASHCOP), Ministry of Health, Tanzania. Due to ethical and legal restrictions, the dataset cannot be publicly shared. However, de-identified data may be made available upon reasonable request to NASCOP through the Ministry of Health, Tanzania (contact) for researchers who meet the criteria for access to confidential data.

## Acknowledgements

I am grateful to God for the gift of life and to KCMC University, particularly the School of Public Health and the Department of Epidemiology and Applied Biostatistics, for offering this essential course. I sincerely thank my supervisor, Dr. Marion Sumari-de Boer, for her consistent support, guidance, and mentorship throughout this research. I also acknowledge Dr. Jegede Feyisayo Ebeneezer for his valuable input and feedback. Special thanks to Mr. Festo Charles for his technical assistance in data extraction and management. Lastly, I appreciate my family, fellow EAB students, and friends for their support and encouragement.

## Author Contribution

**Conceptualization**: Anthony Charles Kavindi, Dr. Marion Sumari-de Boer, and Dr. Jegede Feyisayo Ebeneezer.

**Data curation:** Anthony Charles Kavindi, Festo Charles

**Methodology:** Anthony Charles Kavindi, Dr. Marion Sumari-de Boer, Laura J. Shirima, Deogratius W. Kinoko, Festo Charles, Asteria Karungi Nyongoli, Dr. Jegede Feyisayo Ebeneezer.

**Formal analysis:** Anthony Charles Kavindi

**Writing original draft:** Anthony Charles Kavindi

**Writing-review and editing:** Anthony Charles Kavindi, Dr. Marion Sumari-de Boer, Festo Charles, Dr. Jegede Feyisayo Ebeneezer.

